# Generalizable Model Design for Clinical Event Prediction using Graph Neural Networks

**DOI:** 10.1101/2023.03.22.23287599

**Authors:** Amara Tariq, Gurkiran Kaur, Leon Su, Judy Gichoya, Bhavik Patel, Imon Banerjee

## Abstract

While many machine learning and deep learning-based models for clinical event prediction leverage various data elements from electronic healthcare records such as patient demographics and billing codes, such models face severe challenges when tested outside of their institution of training. These challenges are rooted in differences in patient population characteristics and medical practice patterns of different institutions. We propose a solution to this problem through systematically adaptable design of graph-based convolutional neural networks (GCNN) for clinical event prediction. Our solution relies on unique property of GCNN where data encoded as graph edges is only implicitly used during prediction process and can be adapted after model training without requiring model re-training. Our adaptable GCNN-based prediction models outperformed all comparative models during external validation for two different clinical problems, while supporting multimodal data integration. These results support our hypothesis that carefully designed GCNN-based models can overcome generalization challenges faced by prediction models.

## Introduction

During each patient visit, healthcare centers record the health data of patients in digital systems referred to as Electronic Health Records (EHR) that consist of heterogeneous elements, including demographics, prescriptions, diagnosis, laboratory and radiology test results, encounter notes, procedures, and treatment plans. Structured format of EHR represents data that can take a value within a specified range or from a pre-defined dictionary. Examples of such EHR data include, but are not limited to, medical codes, medications, administrative data, vital signs, and laboratory test outcomes. In the era of digital age, secondary use of structured electronic health records (EHR) for developing machine learning (ML) and deep learning (DL) models for clinical event prediction and digital phenotyping^1,2^ is becoming widely popular and is being clinically adopted for improving the healthcare delivery. However, models trained on a single institution’s data often face severe challenges when applied across multiple different institutions and diverse populations^3^. These challenges usually stem from differences in patient population characteristics such as age, gender, race and common comorbidities among the population, as well as differences in medical practice, manifested in the billing practice of medical procedures and recording and coding of comorbidities as CPT and ICD codes.

ML/DL models commonly leverage ICD and CPT codes to incorporate the clinical status of patients in addition to their demographic features^4,5^. These codes are designed to convert healthcare services to billable revenue. Qualified healthcare coders are responsible for accuracy and completeness of these codes. However, significant differences exist in coding practices between different healthcare institutions^6^. ML/DL models often learn practice patterns of the training institution rather than relevant predictive features and can fail when applied to another institution^7^. Some studies have even shown that time and frequency of lab test order is more important for the model than actually the result of the lab test^8^. A survey paper recently concluded that when tested on external data, more than 20 models trained for prognosis for COVID-19 patients could not outperform univariable predictions made on oxygen saturation level at the time of admission to the hospital, calling into question the utility of ML modeling for clinical event prediction^9^. Proprietary risk prediction models are not exempt from this trend either. Recent studies have shown that the Epic sepsis model achieves subpar performance when validated externally^9^. Models also experience performance decay over time even when deployed in the same institution where it was trained, likely attributed to evolution of population characteristics and practice patterns over time^10^. These challenges limit the generalizability and scalability of ML/DL models that leverage electronic health records for tasks like clinical event prediction or patient phenotyping. Hence, researchers have been motivated to find remedies to the problem of limited generalizability of EHR-based ML/DL models.

A popular applied remedy is the curation of refined clinical features for standardized risk prediction^11^, such as pooled cohort equations^12^. Clinical features are refined to eliminate practice pattern based variations that might arise in the recording of those features. However, curation of these features require extreme manual effort, and hence introduces the possibility of curation errors and is limited to generating smaller datasets. This approach also lacks comprehensiveness and can potentially miss other relevant features that might be predictive for a given DL/ML task contained within the EHR. Such models can only focus on expert-defined clinical features and cannot make use of the vast amount of information available in the electronic health records in general. Research has shown that comprehensive models using a wide variety of EHR outperform models using curated features^13^. Even after valuable curation effort and targeted modeling based on a narrow set of curated features, this approach shows biases among different population groups^12^. Moreover, such models necessitate availability of curated clinical features thus requiring patients to undergo potential tests that may be part of such features. Another approach is harmonizing EHR under standard data models like Observational Medical Outcomes Partnership (OMOP) and Fast Healthcare Interoperability Resources (FHIR). These data models put well-known limitations on granularity of EHR and cannot handle variations in the data patterns themselves^14,15^. These challenges hinder wide-spread adoption of EHR-based ML/DL models across multiple institutions.

Considering the limitations of the previously proposed solutions to the challenge of generalization, we propose a novel solution to adaptability challenges of EHR models through the design of graph-based convolutional neural networks (GCNN). GCNNs have been used to fuse data modalities such as radiological images and demographic information in their two data structures - nodes and edges^1,16^. A few previous studies have hinted towards generalizability properties of GCNN on a small scale for limited patient cohorts^17^. We formalize GCNN model design such that the trained model is generalizable in cross-institutional validation setup and adaptable to the difference between the coded EHR data. In our model design setup, the edge structure of the graph is used to encode patients’ similarity based on structured EHR data elements like ICD and CPT codes. The GCNN models are trained to learn explicitly from data elements selected as node features (e.g., imaging data or selected data elements from EHR) and implicitly from patients’ similarity patterns based on EHR data elements selected for edge formation. These similarity patterns can be systematically adapted to handle temporal and cross-institutional demographic and practice pattern variations while keeping the pre-trained GCNN model applicable as the model only operates on similarity patterns and is agnostic to the exact estimation process used for that similarity pattern.

We validated our generalizable model design framework to solve two clinically relevant problems on completely different populations; 1) prediction of two clinical events for patients hospitalized with positive COVID-19 test: discharge from hospital and mortality using chest X-rays and EHR data elements such as billing codes and demographics features; and 2) prediction of blood transfusion in hospitalized patients using a wide range of EHR data elements (demographic features, CPT and ICD codes, medications, lab tests, vital signs). Hospital discharge and mortality prediction models were trained over data collected from Emory University Healthcare (EUH) network and externally validated on data collected from four geographically disparate sites of Mayo Clinic (MC). For transfusion prediction, data was collected from MC sites of Rochester and Arizona to serve as internal training and testing data while external validation was performed on publicly available MIMIC IV (Medical Information Mart for Intensive Care) dataset that contains data for ICU patients from Beth Israel Deaconess Medical Center in Boston, Massachusetts.

## Methodology

### Graph Convolutional Neural Network

Graph convolutional neural network (GCNN) advanced machine learning by allowing the model designer to choose the definition of ‘*neighborhood*’ to be incorporated by the model through definition of a graph *G*(N, E) where *N* denotes the set of nodes and *E* denotes the set of edges. In this scenario, *i*^th^ sample from the cohort forms *i*^th^ node (***v***_i_) with two feature vectors, i.e., node features (***n***_***i***_) and edge features (***e***_***i***_). An edge between *i*^th^ and *j*^th^ sample, denoted as *ε*_i,j_, is decided based on edge-formation function ϵ(***e***_***i***_, ***e***_***j***_). GCNN model will learn to generate embeddings for *i*^th^ node by manipulating nodes features of this node (***n***_***i***_), and ‘messages’ received from nodes in its edge-connected neighbor (***η***(*i*)). At *k* + 1^th^ graph convolutional layer, the following describes the process of generating embedding of *i*^th^ node (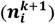)

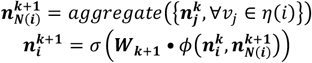

In a supervised leaning scenario where target label for each node is available, node embedding generated by graph convolutional layers is used to predict target label *ŷ*_*i*_ for *i*^th^ node as

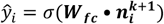

Through backpropagation of loss such as binary cross entropy defined on ground truth *y* and predicted labels *ŷ*, weight matrices ***W***_***k***_∀ *k* ∈ *K* and ***W***_***fc***_ are optimized where the model included *K* graph convolutional layers.

The neighborhood (***η***_i_) of *i*^th^ node ***v***_i_ can be defined based on its edge-connected nodes, i.e., ***η***_i_ = {***v***_j_ ∀*ε*_i,j_ ∈ E }. Messages are sent and received between nodes in a neighborhood. In essence, these messages are features of the nodes (***n***_***j***_, ∀***v***_j_ ∈ ***η***_i_) in the neighborhood (***η***_i_). GCNN model, through its training process, learns the function parameters to manipulate features of the *i*^th^ node (***n***_***i***_) as well as ‘*messages*’ being received through various edge-connected nodes from its neighborhood ***η***_i_. Hence, GCNN is capable of two-fold learning. The model learns from the features of *i*^th^ node (***n***_***i***_) directly, and implicitly learns from information used for edge formation (edge features ***e***_***i***_), through incorporation of ‘*messages*’ from edge-connected nodes (***n***_***j***_, ∀***v***_j_ ∈ ***η***_i_). However, model never directly manipulates edge features (***e***_***i***_).

This is an important advantage of GCNNs as it relates to generalizing a trained model to unseen and diverse populations from external institutes. Since GCNN never has to manipulate edge features (***e***_***i***_) with parametric functions, edge features and edge formation function *ε*(.) can be adapted based on characteristics of the data of the individual institutions when shipping the trained model from one institute to the other. Thus, we based our work upon systematic use of this characteristic of GCNN to build generalizable models using EHR data elements.

#### Adaptable GCNN Design for external use cases

We focused on adaptable edge-formation process to ensure generalizability of our GCNN based models. Trained GCNN model requires consistent formation of node features (***n***_***i***_) in external cohort for its application on external cohort. However, the model does not directly manipulate edge features (***e***_***i***_), and hence, edge formation process *ε*(***e***_***i***_, ***e***_***j***_) can be adapted to suit external cohort without hindering the application of trained GCNN model on external cohort. The following represent two scenarios where such adaptation is crucial.

### Case – 1

Let us assume that edge features of internal cohort are denoted as 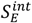 where 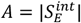 and edge features of the external set are denoted as 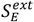 where 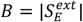 where 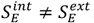. Such distinct feature selection for the two cohorts may be the result of frequency-based selection of common features such as billing codes or administered medication. No existing machine learning model trained on internal feature vectors will be applicable to a separate set of external feature vectors. However, GCNN can tolerate such difference in internal and external cohort by employing these features for edge formation *ε*(.). As explained earlier, GCNN models do no manipulate edge feature vectors directly, but only implicitly use them through ‘messages’ received through these edges.

### Case – 2

Let us assume that edge features for internal and external cohorts are the same, i.e., *S*_E_ with *A* = |*S*_E_|. However, graph formation process is more intelligent than simple thresholding on count-based or binary edge feature vectors for internal 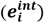 and external 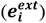 cohorts. For example, edge features may be collected over *T* time intervals, and an edge is formed based on similarity in temporal pattern of these features for nodes *i* and *j*, i.e., 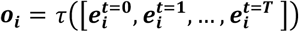 and *ε*(***o***_***i***_, ***o***_***j***_). Even with the same set of features, temporal patterns may be different for internal and external cohorts. For large academic healthcare centers, such patterns may involve both in-patient and out-patient data. For databases collected for critical-care patients only, outpatient data may be missing in temporal patterns. Hence, temporal pattern forming function should be different for internal and external cohort, i.e., *τ*^int^ and *τ*^ext^. Putting limitations on pattern formation function may enable traditional ML models trained on output of internal temporal pattern function *τ*^int^ to be applicable to outputs of external temporal pattern function *τ*^ext^, but graph learning paradigm provides more flexibility. To suit the characteristics of each cohort, *τ*^int^ and *τ*^ext^ may produce output of different dimensions (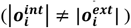), or operate of sequences of different length *T*^int^*and T*^ext^), or even work on different set of features, i.e., 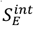 and 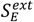 (encompassing the scenario described in case –1).

In terms of patients’ cohort, one node may represent a patient at a certain point in time and edge may denote that two connected nodes/patients are similar in terms of some demographic (e.g., age) or clinical features (e.g., comorbidities). This property has been exploited for detection of Alzheimer and autism spectrum disorder by building graphs with patients as nodes, brain imaging data as node features, and simple demographic features-based similarity used for edge formation^1,16,18,19,20^. We move beyond such modeling by allowing much more comprehensive information to be used as edge features, e.g., all recorded billing codes for patients, or auto-encoder based compressed representation of historical patterns in recorded billing codes and medications.

The primary intuition of our generalizable GCNN design is to represent the measured/recorded health data (e.g. images, lab values) as node features which face minimal chance of variability due to practice pattern, and leverage the adaptability of the edge formation to represent the variable EHR information (e.g. diagnosis and procedure codes).

### Clinical use-cases of the adaptable GCNN design

We validated our model design scheme on two clinically relevant use-cases; 1) prediction of major clinical events (discharge from hospital and mortality) for patients hospitalized with positive RT-PCR test for COVID-19, 2) prediction of need for transfusion for hospitalized patients. Figures 1-a and 1-b show frameworks for both use-cases. The first use-case employs a branched framework where patients marked as highly probable for discharge are evaluated for mortality risk and only in-patient data is used. Second use-case employs historical pattern of recorded procedures, comorbidities, and medication for a patient as well as data recorded during first 48 hours of hospitalization.

**Figure 1.**
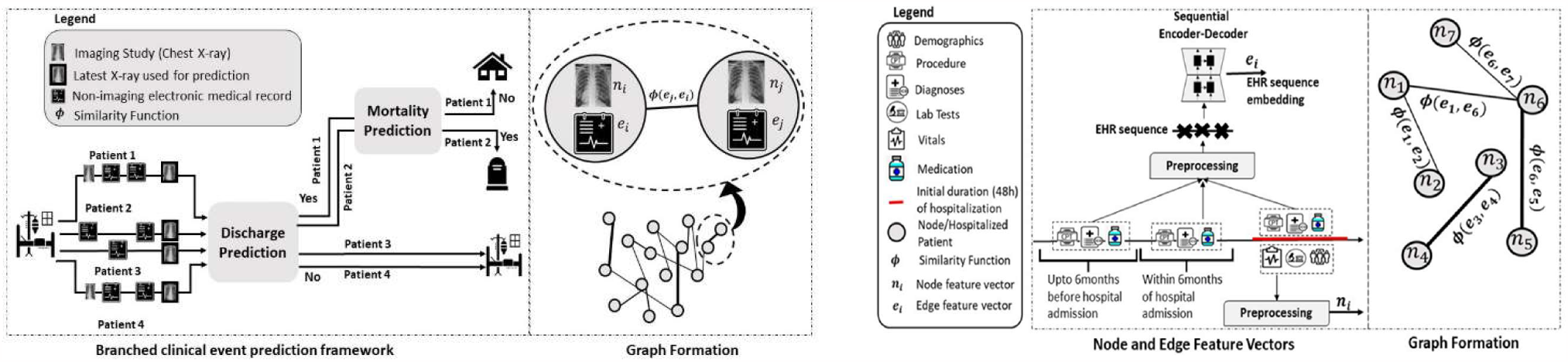
(a) Branched framework and graph formation for use-case 1, (b) Node and edge features processing and graph formation for use-case 2

### Cohort Selection

For use-case 1, internal cohort included all patients admitted to EUH between Jan-Dec 2020 with positive RT-PCR test and for whom chest X-ray examinations (AP view) were acquired at regular intervals during their hospital stay. External cohort was selected with similar criteria from MC (four sites) for the year 2020.

For use-case 2, transfusion prediction model was trained on internal data collected from two sites (Rochester and Arizona) of MC. External validation was performed on two datasets; a) data collected from geographically distant site of MC, i.e., Florida, and b) open-source MIMIC IV dataset. Relatively small number of patients required blood transfusion (approximately 0.5% of hospitalizations in MC in 2019 required blood transfusion). Such a small positivity rate of transfusion drove us to curate a training dataset through propensity matching for the control group.

Demographic features as well as 5 major comorbidities groups including metabolic disorders, hypertensive disease, heart disease, acute kidney failure adverse effects of drugs, were used as confounders to perform one-on-one propensity matching with cases (hospitalizations with blood transfusion) to select control groups (hospitalizations without blood transfusion). Case and control groups were perfectly balanced in terms of confounding variables in our propensity matched training data. Salient characteristics of all cohorts are described in Table 1.

**Table 1.** Cohort characteristics - Race X: American Indian/Alaskan Native, Race Y: Native Hawaiian/Pacific Islander, Ethnicity Z: Hispanic or Latino

|  |  | COVID-19 Clinical Event Prediction Cohort |  |  |  | Transfusion Prediction Cohort |  |  |  |  |
| --- | --- | --- | --- | --- | --- | --- | --- | --- | --- | --- |
|  | Split (Patients, Hospitalization) | Train (1578, 5741) | Validation (184, 527) | Test (439, 1545) | External test (1082, 3800) | Train (8752, 11290) | Validation (993, 1286) | Test (2472, 3151) | Internal Hold-out site (3041, 4042) | External test (68,68) |
| Age | mean+/-std | 58.9 +/- 17.5 | 59.4 +/- 18.0 | 61.0 +/- 17.1 | 65.3 +/- 15.4 | 62.1 +/- 18.2 | 62.6 +/- 19.2 | 62.3 +/- 18.3 | 63.5 +/- 14.6 | 69.0 +/- 15.3 |
| Sex | Female | 785(49.7%) | 90(48.9%) | 215(49.0%) | 400(37.0%) | 3989(45.6%) | 438(44.1%) | 1141(46.2%) | 1459(48.0%) | 26(38.2%) |
|  | Male | 793(50.3%) | 94(51.1%) | 224(51.0%) | 682(63.0%) | 4763(54.4%) | 555(55.9%) | 1331(53.8%) | 1582(52.0%) | 42(61.8%) |
| Race | White | 384(24.3%) | 43(23.4%) | 127(28.9%) | 892(82.4%) | 7688(87.8%) | 869(87.5%) | 2175(88.0%) | 2332(76.7%) | 48(70.6%) |
|  | Black | 1014(64.3%) | 121(65.8%) | 270(61.5%) | 86(7.9%) | 291(3.3%) | 33(3.3%) | 91(3.7%) | 455(15.0%) | 8(11.8%) |
|  | Asian | 36(2.3%) | 6(3.3%) | 9(2.1%) | 46(4.3%) | 207(2.4%) | 20(2.0%) | 59(2.4%) | 101(3.3%) | 0(0%) |
|  | Race X | 7(0.4%) | 0(0%) | 4(0.9%) | 14(1.3%) | 126(1.4%) | 13(1.3%) | 32(1.3%) | 10(0.3%) | 0(0%) |
|  | Race Y | 4(0.3%) | 0(0%) | 2(0.5%) | 2(0.2%) | 23(0.3%) | 0(0%) | 9(0.4%) | 9(0.3%) | 0(0%) |
|  | Unknown | 133(8.4%) | 14(7.6%) | 27(6.2%) | 42(3.9%) | 417(4.8%) | 58(5.8%) | 106(4.3%) | 134(4.4%) | 12(17.6%) |
| Ethnicity | Not Z | 1362(86.3%) | 155(84.2%) | 377(85.9%) | 971(89.7%) | 8036(91.8%) | 896(90.2%) | 2276(92.1%) | 2767(91.0%) | 56(82.4%) |
|  | Z | 114(7.2%) | 19(10.3%) | 30(6.8%) | 99(9.1%) | 421(4.8%) | 62(6.2%) | 112(4.5%) | 167(5.5%) | 0(0%) |
|  | Unknown | 102(6.5) | 10(5.4%) | 32(7.3%) | 12(1.1%) | 295(3.4%) | 35(3.5%) | 84(3.4%) | 107(3.5%) | 12(17.6%) |
| Comorbidities | Diabetes | 746(47.3%) | 84(45.7%) | 205(46.7%) | 409(37.8%) | 2533(28.9%) | 268(27.0%) | 713(28.8%) | 1050(34.5%) | 0(0.0%) |
|  | Hypertension | 1076(68.2%) | 119(64.7%) | 311(70.8%) | 711(65.7%) | 5508(62.9%) | 620(62.4%) | 1575(63.7%) | 2176(71.6%) | 48(70.6%) |
|  | Heart Disease | 831(52.7%) | 83(45.1%) | 230(52.4%) | 814(75.2%) | 5705(65.2%) | 659(66.4%) | 1647(66.6%) | 1925(63.3%) | 0(0.0%) |
|  | Kidney Disease | 266(16.9%) | 22(12.0%) | 79(18.0%) | 554(51.2%) | 3932(44.9%) | 449(45.2%) | 1164(47.1%) | 1526(50.2%) | 0(0.0%) |

### Model Design

Figure 2 shows distributions of subgroups of billing code sets (CPT and ICD) for use-case 1 from two different institutions. External institute used a much larger number diagnostic tests indicated by higher bars for diagnostic radiology and drug assay subgroups. While blood disease seems to be more common among patients in internal institute than in external institute, external cohort had a larger fraction of patients with metabolic disorders and heart disease. Such data elements require adaptation when experimenting from external cohort, and hence are suitable for edge feature formation.

**Figure 2.**
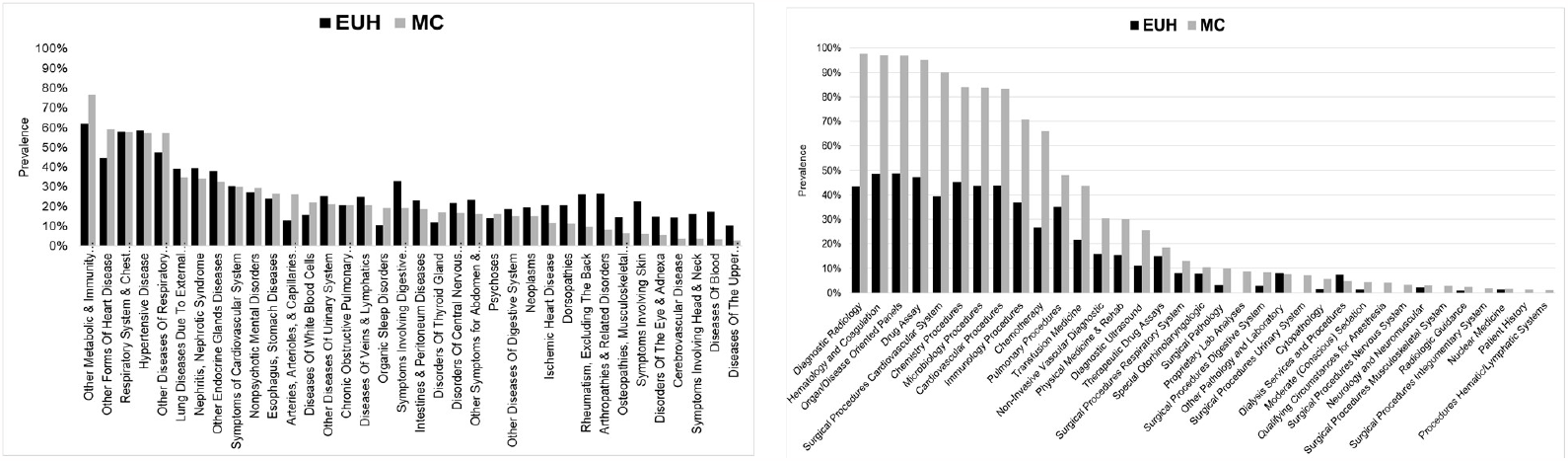
Billing codes distribution; a) ICD, b) CPT, in internal and external sets for COVID-19 patients’ cohorts

Such variations are handled by our generalizable model design which relies on unique adaptable learning paradigm of GCNN model as described earlier. Chest X-rays and tabular data (demographics, and CPT and ICD codes) were available for case-study 1. Image features extracted from pre-trained DenseNet-121 models were used as node features (***n***_***i***_). All tabular data elements were evaluated as edge feature vectors (***e***_***i***_) for effective edge formation. CPT and ICD codes were mapped to their corresponding subgroups in CPT and ICD code hierarchies respectively, and finally represented as one-hot feature vectors.

Transfusion prediction model employed a larger variety of EHR features. Latest results of selected labs (recorded as Normal/Abnormal/Unknown), trend of change (gradient) in five important vital signs (temperature, mean arterial pressure (MAP), SpO2, pain score, pulse rate) recorded during the first 48 hours of hospitalization, demographic features, and free-text field of reason for visit vectorized under tf-idf featurization scheme were concatenated to form node features for this model. Edge features (*e*_i_) were generated by a temporal embedding model *τ*(.) for variable data elements like billing codes (CPT and ICD) and medication.

### Temporal Embedding Model

Embedding model *τ*(.) encodes temporal patterns recorded as three-point sequences; Timepoint 1 (T1): data collected between 6 and 12 months before hospitalization, Timepoint 2 (T2): data collected between 6 months before hospitalization to the time of hospitalization, Timepoint 3 (T3): data collected within first 48 hours of hospitalization.

Temporal embedding model τ(.) is essentially an LSTM based encoder-decoder architecture. The feature vector at each time point is encoded to a latent space such that when decoded, it generates the feature vector of the next time point. Hence, the model is trained in a self-supervised fashion with no regard to any downstream prediction label. Once the model has been trained, the last hidden state vector generated in response to an input sequence can be used as an embedded edge feature vector *e*_i_ encompassing temporal information encoded in the data elements used as input. Our transfusion prediction model operates on a graph of patients where edges between patients are decided based on similarity in their embedded vectors *e*_i_. As explained in Methodology section, this temporal embedding model is trained separately for external data, while keeping the GCNN-based transfusion prediction model trained on internal cohort applicable over external cohort.

Three timepoints sequence design was selected experimentally as further finer-grained splitting of historical data resulted in many empty timepoints (time interval with no available data). This is due to the nature of data collected as inpatient vs outpatient. Most historical data is outpatient data except for cases where a patient was hospitalized in the last one year as well. Outpatient data is much more sparse than inpatient data. Note that external data was collected from MIMIC IV which is very different from the internal cohort. MIMIC data records patient hospitalizations with no outpatient data. Hence historical data is only available if the patient was hospitalized within the last one year as well. Still, retraining the embedding model provided a chance for adaptation to such a different scenario.

## Results

We performed thorough experimental evaluation for both use-cases. As both use-cases involve multiple data elements (including EHR data elements and chest X-rays) and the proposed approach is based on fusion of all data elements through GCNN-based models, an intuitive comparative baseline is formed by single-modality models, each using only one of the data elements. Therefore, we trained and evaluated such single modality models for all three clinical events. In addition, we also employed a traditional fusion modeling approach, i.e., late fusion, which gathered target label probability estimates from single modality models and processed them together through a meta-learner for final target prediction. Tables 2 and 3 show performance of clinical event predictors for use-cases 1 and 2, respectively, for both internal held-out test sets and external sets.

**Table 2.** Performance of clinical event prediction models for COVID-19 patients

| Model | Internal |  |  | External |  |  |
| --- | --- | --- | --- | --- | --- | --- |
|  | Sensitivity | Specificity | AUROC | Sensitivity | Specificity | AUROC |
|  | Hospital Discharge Prediction |  |  |  |  |  |
| Non-imaging (EHR) | 72.9 [71.7-74.1] | 59.9 [58.2-61.5] | 71.5 [70.4-72.5] | 48.9 [48.2-49.7] | 64.1 [62.8-65.4] | 57.8 [56.9-58.7] |
| Images (X-rays) | 71.7 [70.5-72.9] | 65.9 [64.2-67.4] | 74.9 [73.8-76.0] | 59.6 [58.9-60.3] | 54.6 [53.4-56.0] | 60.0 [59.1-60.9] |
| Late fusion | 69.7 [68.4-70.8] | 65.5 [64.1-67.2] | 74.5 [73.4-75.6] | 51.2 [50.4-51.9] | 61.3 [60.0-62.7] | 58.1 [57.2-59.0] |
| GCNN-Demo | 70.2 [69.0-71.4] | 68.9 [67.3-70.5] | 76.0 [75.0-77.0] | 60.7 [60.0-61.4] | 62.2 [60.9-63.5] | 65.2 [64.3-66.1] |
| GCNN-CPT | 71.1 [69.9-72.3] | 69.6 [68.2-71.4] | 77.1 [76.1-78.2] | 64.8 [64.1-65.4] | 57.4 [55.9-58.8] | 64.6 [63.7-65.5] |
| GCNN-ICD | 69.5 [68.2-70.6] | 70.2 [68.7-71.8] | 76.2 [75.1-77.3] | 64.5 [63.7-65.2] | 66.3 [65.1-67.5] | 70.0 [68.7-70.4] |
|  | Mortality Prediction |  |  |  |  |  |
|  | Sensitivity | Specificity | AUROC | Sensitivity | Specificity | AUROC |
| Non-imaging (EHR) | 86.4 [84.2-89.4] | 83.8 [82.5-85.3] | 86.7 [84.9-88.5] | 79.3 [77.2-81.5] | 81.4 [80.3-82.6] | 86.7 [85.7-87.7] |
| Images (X-rays) | 86.4 [84.0-89.0] | 77.8 [76.4-79.5] | 88.1 [86.9-89.3] | 64.9 [62.3-67.5] | 35.6 [34.2-37.1] | 48.7 [46.9-50.4] |
| Late fusion | 85.6 [83.0-88.5] | 81.1 [79.8-82.8] | 88.6 [87.2-90.2] | 76.0 [74.0-78.3] | 82.2 [81.1-83.4] | 81.6 [80.3-82.9] |
| GCNN-Demo | 84.7 [82.3-87.6] | 81.1 [79.7-82.6] | 89.4 [88.4-90.7] | 78.1 [76.1-80.2] | 86.1 [85.1-87.2] | 88.8 [87.9-89.8] |
| GCNN-CPT | 74.6 [71.3-78.0] | 84.5 [83.0-85.9] | 86.6 [85.2-88.0] | 83.1 [81.3-84.8] | 86.1 [85.2-87.4] | 91.4 [90.6-92.3] |
| GCNN-ICD | 84.7 [82.3-87.9] | 82.5 [81.1-83.9] | 90.1 [89.0-91.3] | 81.4 [79.6-83.5] | 74.6 [73.2-76.0] | 85.3 [84.2-86.5] |

**Table 3.** Performance of all models for prediction of transfusion for hospitalized patients; ‘--’ were added to the places where the performance cannot be computed due to missing data.

| Model | Internal |  |  | Held-out site |  |  | External |  |  |
| --- | --- | --- | --- | --- | --- | --- | --- | --- | --- |
|  | Sensitivity | Specificity | AUROC | Sensitivity | Specificity | AUROC | Sensitivity | Specificity | AUROC |
| **Demographics** | 54.0<br>[52.0-56.0] | 51.5<br>[50.0-52.9] | 54.2<br>[52.8-55.6] | 53.8<br>[52.2-55.7] | 45.7<br>[44.3-47.0] | 50.5<br>[49.3-51.9] | 60.0<br>[50.0-75.0] | 66.7<br>[63.2-70.2] | 63.3<br>[54.2-76.3] |
| **CPT** | 54.8<br>[52.8-56.8] | 66.1<br>[64.8-67.4] | 61.4<br>[59.9-62.9] | 57.0<br>[55.3-58.7] | 61.6<br>[60.3-62.7] | 59.7<br>[58.4-60.9] | -- | -- | -- |
| **ICD** | 59.0<br>[56.9-60.9] | 63.0<br>[61.7-64.4] | 62.3<br>[60.9-63.7] | 62.0<br>[60.3-63.7] | 63.3<br>[62.2-64.6] | 63.7<br>[62.5-64.9] | 60.0<br>[50.0-75.0] | **71.4**<br>[68.4-75.5] | 58.6<br>[52.2-66.5] |
| **Lab Test** | 59.8<br>[57.8-61.7] | 62.3<br>[60.9-63.6] | 64.5<br>[63.1-65.7] | 50.5<br>[48.6-52.2] | 59.0<br>[57.9-60.3] | 56.3<br>[54.9-57.4] | 60.0<br>[50.0-75.0] | 49.2<br>[45.3-52.8] | 45.1<br>[34.2-54.2] |
| **Medications** | 58.1<br>[56.2-59.9] | 62.9<br>[61.5-64.3] | 63.2<br>[61.7-64.6] | 57.0<br>[55.1-58.7] | 61.8<br>[60.6-63.0] | 61.1<br>[59.8-62.3] | 60.0<br>[50.0-75.0] | 69.8<br>[66.7-73.7] | 61.0<br>[50.0-76.3] |
| **Reason for visit** | 35.2<br>[33.4-37.2] | **79.6**<br>[78.5-80.8] | 58.2<br>[57.0-59.7] | 31.1<br>[29.4-32.7] | **77.0**<br>[76.1-78.1] | 55.5<br>[54.3-56.6] | -- | -- | -- |
| **Vitals** | 61.6<br>[59.7-63.4] | 58.0<br>[56.6-59.3] | 62.8<br>[61.4-64.1] | 60.9<br>[59.3-62.7] | 54.5<br>[53.2-55.7] | 59.4<br>[58.1-60.6] | 80.0<br>[75.0-100.0] | 31.7<br>[28.1-35.2] | 43.2<br>[32.0-49.2] |
| **Late Fusion** | 64.0<br>[62.1-66.0] | 64.2<br>[62.9-65.5] | 69.9<br>[68.6-71.2] | 68.0<br>[66.4-69.4] | 60.9<br>[59.8-62.2] | 68.3<br>[67.2-69.4] | 60.0<br>[50.0-75.0] | 54.0<br>[50.8-57.9] | 44.4<br>[31.3-56.0] |
| **GNN** | **73.8**<br>[72.0-75.6] | 65.4<br>[64.1-66.7] | **77.4**<br>[76.3-78.5] | **70.2**<br>[68.6-71.7] | 70.0<br>[68.9-71.2] | **77.1**<br>[76.1-78.1] | **80.0**<br>[66.0-95.0] | 69.8<br>[66.7-73.6] | **70.8**<br>[62.5-84.7] |

For use case 1, other than the obvious difference in coding patterns (Figure 2), the patient populations are significantly different between EUH and MC (internal and external institutions, respectively) - (1) 49% female patients in EUH while 37% female in MC; (2) 61.5% African American in EUH and 7.9% in MC; (3) as comorbidities, 16.9% kidney disease in EUH and 51.2% in MC; 52.4% cardiovascular disease in EUH while 75.2% in MC (Table 1). For use-case 2, the external cohort was significantly small (68 patients); however similar variations in patient population were also observed.

In the challenging scenario formed by vast differences in internal and external datasets, individual modality classifiers and traditional fusion models struggle when presented with external data. On the other hand, GNN based models tend to fare better under similar settings. For use-case 1, late fusion model achieved 0.58 [0.52 - 0.59] AUROC on the external dataset for hospital discharge prediction while the GCNN achieved 0.70 [0.68-0.70]. For mortality prediction, late fusion model achieved 0.81[0.80-0.82] AUROC on external dataset while the GCNN model achieved 0.91 [0.90 - 0.92]. For the use-case 2, we observed more gaps in performance due to missing/incomplete data in the external dataset - late fusion achieved 0.44[0.31-0.56] while the GCNN model achieved 0.7 [0.62-0.84].

We hypothesize that superior performance of GCNN on external data is due to the adaptation of the edge formation function. To test this hypothesis, we applied GCNN based models without adaptation of edge formation function on external for all our clinical event prediction tasks and compared results with application of GCNN based models with edge formation function adaptation. The models suffer significant performance loss in majority of the cases when used without edge formation function adaptation (Table 4). Hence, we can safely conclude that the generalization and adaptability capabilities of GCNN based models arise from its unique ability to adapt edge formation process to suit new population even after model training.

**Table 4.** Performance of GCNN models on external cohorts with and without edge adaptation on external cohorts

| Model | With adaptation |  |  | Without adaptation |  |  |
| --- | --- | --- | --- | --- | --- | --- |
|  | Sensitivity | Specificity | AUROC | Sensitivity | Specificity | AUROC |
| **Hospital Discharge Prediction** |  |  |  |  |  |  |
| **GCNN-CPT** | 64.8 [64.1-65.4] | 57.4 [55.9-58.8] | 64.6 [63.7-65.5] | 63.4 [62.7-64.1] | 58.0 [56.5-59.4] | 64.2 [63.4-65.1] |
| **GCNN-ICD** | 64.5 [63.7-65.2] | 66.3 [65.1-67.5] | 70.0 [68.7-70.4] | 63.3 [62.6-64.0] | 61.3 [59.9-62.6] | 65.8 [65.0-66.7] |
| **Mortality prediction** |  |  |  |  |  |  |
| **GCNN-CPT** | 83.1 [81.3-84.8] | 86.1 [85.2-87.4] | 91.4 [90.6-92.3] | 69.4 [67.2-71.7] | 74.4 [73.1-75.8] | 79.6 [78.3-80.9] |
| **GCNN-ICD** | 81.4 [79.6-83.5] | 74.6 [73.2-76.0] | 85.3 [84.2-86.5] | 73.6 [71.4-76.0] | 74.9 [73.4-76.2] | 82.3 [81.0-83.5] |
| **Transfusion Prediction** |  |  |  |  |  |  |
| **GCNN** | 80.0 [66.7-100.0] | 69.8 [65.4-74.6] | 70.8 [57.9-85.6] | 60.0 [33.3-75.0] | 74.6 [70.4-79.2] | 63.2 [49.4-76.8] |

## Discussion

As highlighted in the literature^7,8^, the challenges related to generalization limit the application scope of ML/DL models that could otherwise leverage the rich electronic health records for tasks such as clinical event prediction or patient phenotyping. In this study, we propose an adaptable GCNN framework for EHR modeling that can be easily generalizable across institutions where the difference in patient population and coding practices are significant. The GCNN framework allows to choose the definition of patient/case similarity to be incorporated into the model through definition of a graph which not only mimics the parts of clinical decision making but can also be utilized to overcome the generalizability limitation of traditional ML/DL models. While the GCNN model learns from the features of node directly, it implicitly learns from information used for edge formation through incorporation of edge-connected nodes. Thus, our generalizable GCNN design primarily represents the measured/recorded health data (e.g. images, lab values) as node features which usually have minimal variability across sites, and leverage the adaptability of the edge formation to represent the variable EHR information (e.g. diagnosis and procedure codes).

We validated the proposed generalizable GCNN model design framework to solve two important clinical use-cases; 1) prediction of adverse clinical events for COVID-19 patients, and 2) prediction of blood transfusion in hospitalized patients. We trained the GCNN models using data from one institution (EUH/MC) and validated externally on MC and publicly available MIMICIV datasets, respectively. During our experimentations, even though the performances on the internal datasets were close, GCNN models consistently outperformed the traditional ML/DL models on the external datasets. We hypothesized that this performance trend is due to the ability of graph-based models to adapt their edge formation functions without requiring any re-training or fine tuning the model itself.

To our knowledge, we are the first to report the edge adaptable GCNN property to improve the generalizability of ML/DL model in healthcare settings. Our proposed design has several important advantages. First, the model trained on an internal dataset does not need fine-tuning or retraining on the external data, even when the EHR data structure and coding frequency differs significantly between the institutions. Second, the graph design implicitly models the similarity between the patients and thus mimics the clinical decision making. Third, the adaptable edge formation technique allows to explore institution specific variables to define the patient/case similarity and provide flexible design choice. Fourth, graph design allows integration of multi-modal data (images + EHR).

There are several limitations in this study, such as those associated with a retrospective design of both use-cases. In addition, for the MIMIC dataset, the timestamp associated with the CPT code and reason for admission were missing, thus we were not able to evaluate the model performance using those data elements. Given the low prevalence, we used propensity score matching to select the control cases which provided only a selective sample for validation.

## Conclusion

We proposed a novel solution to the challenges faced by machine learning and deep learning-based models relying on electronic health records for predictive modeling for patient populations. Generally, such models suffer from poor generalization capabilities due to differences in medical practice patterns and patient population characteristics when applied outside of their institute of training. Our systematic design of graph based convolutional neural networks implicitly learns from such varying data elements of electronic health records through their use in the edge formation process. Edge formation function can be adapted to suit the new population when models are to be tested externally without needing any retraining of the originally trained model. We proved the benefits of our approach through its application on two clinically relevant problems; each tested on two diverse populations. We included a wide variety of electronic health records data elements as well as imaging information, indicating that our approach is capable of handling complex multi-modal data while developing highly adaptable models.

## Data Availability

Data collected from Emory University Healthcare network and Mayo Clinic is confidential, and can only be made available through request to these institutions.

